# *Mycobacterium tuberculosis*-specific T cell activation identifies individuals at high risk of tuberculosis disease

**DOI:** 10.1101/2020.06.26.20135665

**Authors:** Cheleka A.M. Mpande, Munyaradzi Musvosvi, Virginie Rozot, Boitumelo Mosito, Timothy D. Reid, Constance Schreuder, Tessa Lloyd, Nicole Bilek, Huang Huang, Gerlinde Obermoser, Mark M. Davis, Morten Ruhwald, Mark Hatherill, Thomas J. Scriba, Elisa Nemes, ACS Study Team

## Abstract

**Background:** Provision of tuberculosis preventive treatment (TPT) to individuals with *Mycobacterium tuberculosis* (M.tb) infection (TBI) is a key strategy to reduce the global tuberculosis burden. Tuberculosis risk is significantly higher after recent compared to remote TBI. We aimed to define a blood-based biomarker, measured with a simple flow cytometry assay, to stratify different stages of TBI to infer risk of disease.

**Methods:** Healthy adolescents were serially tested with QuantiFERON-TB Gold (QFT) to define recent (QFT conversion <6 months) and remote (persistent QFT+ for >1 year) TBI. M.tb-specific T cells were defined as IFN-g+TNF+CD3+ cells upon CFP-10/ESAT-6 or M.tb lysate stimulation. ΔHLA-DR median fluorescence intensity (MFI) was defined as the difference in HLA-DR expression between M.tb-specific and total T cells. Biomarker performance was assessed by blinded prediction in untouched test cohorts with recent *versus* remote TBI or tuberculosis disease, and unblinded analysis of asymptomatic adolescents with TBI who remained healthy (non-progressors) or who progressed to microbiologically-confirmed disease (progressors).

**Findings:** In the test cohorts, frequencies of M.tb-specific T cells differentiated between QFT- (n=25) and QFT+ (n=47) individuals [area under the ROC curve (AUCROC): 0.94; 95%CI: 0.87-1.00]. ΔHLA-DR MFI significantly discriminated between recent (n=20) and remote (n=22) TBI (AUCROC 0.91; 95%CI: 0.83-1.00); remote TBI and newly diagnosed tuberculosis (n=19; AUCROC 0.99; 95%CI: 0.96-1.00); and between tuberculosis progressors (n=22) and non-progressors (n=34; AUCROC 0.75, 95%CI: 0.63-0.87).

**Interpretation:** The ΔHLA-DR MFI biomarker can identify individuals with recent TBI and those with disease progression, allowing targeted provision of TPT to those at highest risk of tuberculosis.

## Introduction

Tuberculosis is an ongoing global threat causing an estimated ten million incident cases and approximately 1.5 million deaths in 2018 (1), and is the leading cause of death due to a single infectious agent. A key strategy to alleviate the global burden of tuberculosis, promoted by the World Health Organisation, is to provide tuberculosis preventive therapy (TPT) to individuals with *Mycobacterium tuberculosis* (M.tb) infection (TBI), who are at risk of disease progression (2). Current diagnostic tests can identify persons with tuberculosis (sputum-based microbiological tests) and those with TBI [tuberculin skin test (TST) or IFN-γ release assays (IGRAs)]. However, these tests cannot distinguish who, among those with asymptomatic TBI are at high risk of progressing to active disease and should receive TPT.

In countries with low tuberculosis burden (<100 cases/1 million people per year) and/or high income, TPT is standard of care for persons with clinical and epidemiological risk factors or diagnosis of TBI, and reduces the risk of tuberculosis (3, 4). On the other hand, in many countries where M.tb is endemic more than 50% of adults may have TBI (5), making provision of TPT infeasible and unaffordable, especially without knowledge of exposure history. Provision of TPT to all M.tb-infected individuals is also not universally practiced because risk of reinfection after treatment completion is high in high transmission settings (6, 7). Furthermore, given the small proportion of M.tb infected individuals who are actually at risk of tuberculosis progression, provision of TPT to all those infected exposes many individuals to unnecessary side-effects.

The highest risk of tuberculosis progression occurs within the first 1-2 years after primary infection (8-10). By contrast, established, remote infection is associated with a much lower risk of tuberculosis (8, 9). In fact, remote TBI has been associated with “protective immunity” against disease progression (11, 12), and there is evidence that many individuals with remote M.tb exposure may actually have cleared M.tb (13). A blood-based biomarker that could distinguish between recent and remote infection would therefore allow targeted TPT to those with recent infection and could potentially transform the clinical management of tuberculosis, regardless of setting (14). Unfortunately, current diagnostic tests for TBI measure immunological sensitization to M.tb and cannot distinguish between recent and remote TBI. Improving tools for detecting TBI and testing for progression to tuberculosis is a key priority in the Global Plan to end TB (15).

M.tb-specific T cell features such as activation (16-19), differentiation (20, 21) and polyfunctional profiles (22, 23) have shown promising diagnostic applications to distinguish TBI from tuberculosis, and to monitor treatment response. The underlying hypothesis is that all these biomarkers reflect M.tb antigen load, which is higher during tuberculosis compared to controlled TBI, and that tuberculosis treatment effectively reduces M.tb load. We postulated that recent TBI is associated with a high initial M.tb load that is ultimately controlled by immune responses in those with remote infection, who do not progress to primary disease. We also postulated that progression from TBI to disease is associated with a gradual increase in M.tb load (24). This is supported by our previous study in progressors, which showed increased inflammation and activation of bulk CD4 T cells prior to clinical onset (25). In line with this principle, Halliday and colleagues discovered that proportions of M.tb-specific TNF+IFN-γ-IL-2-CD4 T cells with an effector (TEFF) phenotype distinguished between recent and remote TBI (26).

In this study, we hypothesised that recent TBI and progression to tuberculosis are associated with higher levels of M.tb-specific T cell activation compared to remote TBI when risk of disease progression is low. We aimed to develop a simplified assay that could stratify, for the first time, the entire spectrum of M.tb from acquisition of infection to clinical disease and through treatment response, and that is amenable to clinical translation and further large-scale clinical validation.

## Methods

### Study Design

We designed a retrospective study in adolescents and adults to compare immune responses between recent and remote TBI, TBI and tuberculosis disease, as well as between tuberculosis progressors and non-progressors using cryopreserved peripheral blood mononuclear cells (PBMC).

### Participants

Adolescent and adult participants were enrolled in observational studies approved by the University of Cape Town Human Research Ethics Committee (protocol references: 045/2005, 088/2008, 102/2017).

#### Adolescents

Different groups of adolescents were selected (Figure 1) from a large epidemiological study conducted in the greater Worcester area, Western Cape, South Africa, from July 2005 through February 2009 (10). Healthy 12-18 years old participants were enrolled at high schools; those pregnant or lactating, and those who reported acute or chronic medical conditions resulting in hospitalization within 6 months prior to enrolment were excluded. HIV testing was allowed only in participants diagnosed with tuberculosis during follow-up, and those who were HIV-positive were excluded from this study. Adolescent and parents or legal guardians provided written, informed assent and consent, respectively. QuantiFERON-TB Gold In-Tube (QFT; Cellestis) testing was performed to measure TBI and PBMC were collected at enrolment and at 6-monthly intervals during a 2-year follow-up in a subset of the cohort. Participants with tuberculosis symptoms or household contact with a tuberculosis patient were investigated for disease throughout follow-up.

**Figure 1:**
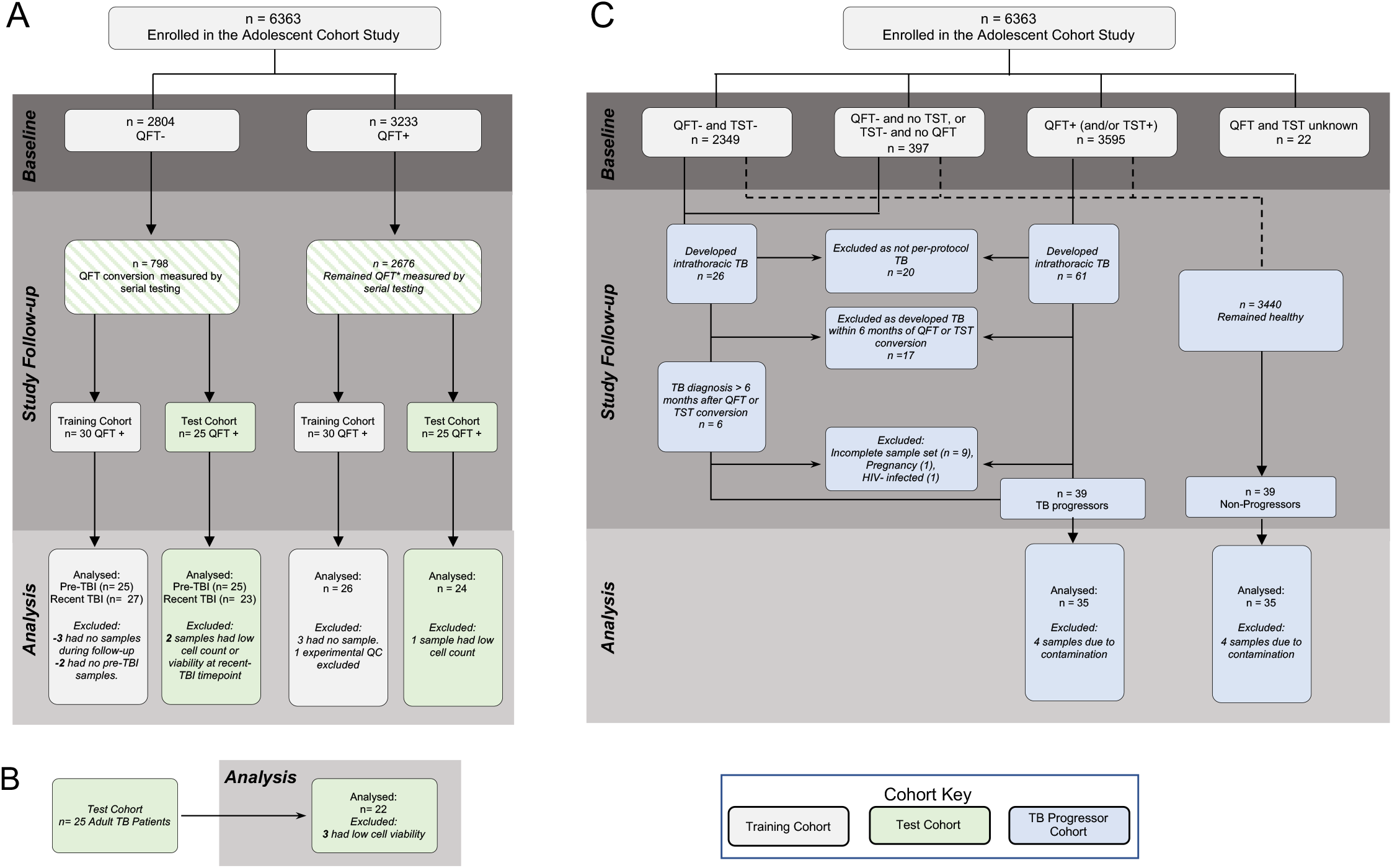
Study Consort. Adolescent participants were selected based on PBMC sample availability from a larger epidemiological study, the Adolescent Cohort Study (10). (A) Inclusions and exclusions for the recent and remote TBI training and test cohorts. (B) Adults with tuberculosis diagnosis sampled cross-sectionally (31). (C) Reasons for inclusion and exclusion of participants as progressors and non-progressors cohort. QFT = QuantiFERON-TB Gold; TST = tuberculin skin test.

### Definition of recent and remote TBI

Recent TBI was defined in healthy participants by two negative QFT tests (IFN-g<0.35 IU/mL; assay performed and interpreted according to the manufacturer’s specifications) followed by two positive QFT tests (IFN-g≥0.35 IU/mL) 6 months apart over 1.5 years. Remote TBI was defined in healthy participants by 4 consecutive QFT positive tests 6 months apart over 1.5 years. Here we included results from the first QFT- and the first QFT+ visit (1 year apart) for participants with recent TBI, and the third or fourth QFT+ visit for participants with remote TBI. Adolescents selected based on PBMC availability were split into a training cohort and a blinded test cohort (Figure 1A, Supplementary Table 1).

### Definition of progressors and non-progressors

Adolescents with TBI, defined by a positive QFT test and/or a positive TST (induration >10mm; 0.1 mL dose of purified protein derivative RT-23, 2-TU, Statens Serum Institut), who developed microbiologically-confirmed, intrathoracic tuberculosis during the 2-year follow-up were included as progressors, while those who remained healthy were included as non-progressors (Figure 1C). Individuals included here were a sub-group of adolescents studied to identify a transcriptomic blood signature for tuberculosis risk, selected based on sample availability (25, 27). Tuberculosis was diagnosed by either two sputum samples positive on smear microscopy for acid-fast bacilli or one sputum sample positive for *M*.*tb* by liquid culture (27). Only samples collected before tuberculosis diagnosis were analysed.

#### Adults

Patients above 18 years old with recently diagnosed tuberculosis (positive XpertMTB/RIF, Cepheid), who provided written informed consent were recruited from the greater Worcester area clinics, Western Cape, South Africa, from January through August 2015. All adults with HIV-infection, anaemia (Haemoglobin <8.0d/dL), body mass index ≤17, with signs of other chronic illnesses that were not consistent with tuberculosis and those who had already started tuberculosis treatment were excluded from the study.

### PBMC stimulation and staining protocols

Cryopreserved PBMC were thawed and stimulated with peptide pools spanning full length CFP-10/ESAT-6, antigens used in the QFT assay, for 18 hours (tuberculosis, recent and remote TBI groups) or M.tb lysate (H37Rv) for 12 hours (progressors and non-progressors). Cells were stained and analysed by flow cytometry (Supplementary Tables 2, 3, and 4). M.tb-specific cells were defined by cytokine expression, detected either by intra-cellular cytokine staining (ICS; tuberculosis, recent and remote TBI groups) or gene expression from sorted, single CD69+CD137+ and/or CD69+CD154+ cells (progressors and non-progressors). Detailed information is provided in the Supplementary Methods.

### Data Analysis

#### Responder definition

We identified responders as individuals with relative counts of IFN-γ+TNF+ CD3+ T cells in the antigen-stimulated condition that were significantly higher (p≤0.01 by Fisher’s Exact test) than the unstimulated condition, and had a fold change of antigen-specific T cell frequencies of ≥3.

#### Progressor and non-progressors transcriptomic analysis

TCRαβ sequences were amplified using a panel of TCRαβ primers and further amplified in a nested PCR before sequencing on a MiSeq (Illumina) instrument, as described previously (28, 29). Expression of 20 mRNA transcripts was detected within each sorted T cell, as described previously (28, 29). Co-expression of IFNG+TNF+ was defined by 5 or more IFNG and TNF reads per cell. Only samples with 10 or more IFNG and TNF co-expressing cells were analysed for HLA-DR expression (see below).

#### T cell activation biomarker definition

In the training and progressor cohorts, we explored different calculations to define the T cell activation biomarker: HLA-DR+ T cells as a percentage of total IFN-γ+TNF+ T cells, HLA-DR median fluorescent intensity (MFI) ratio (Equation 1) and Delta (Δ) HLA-DR MFI (Equation 2, Figure 2D).

**Figure 2.**
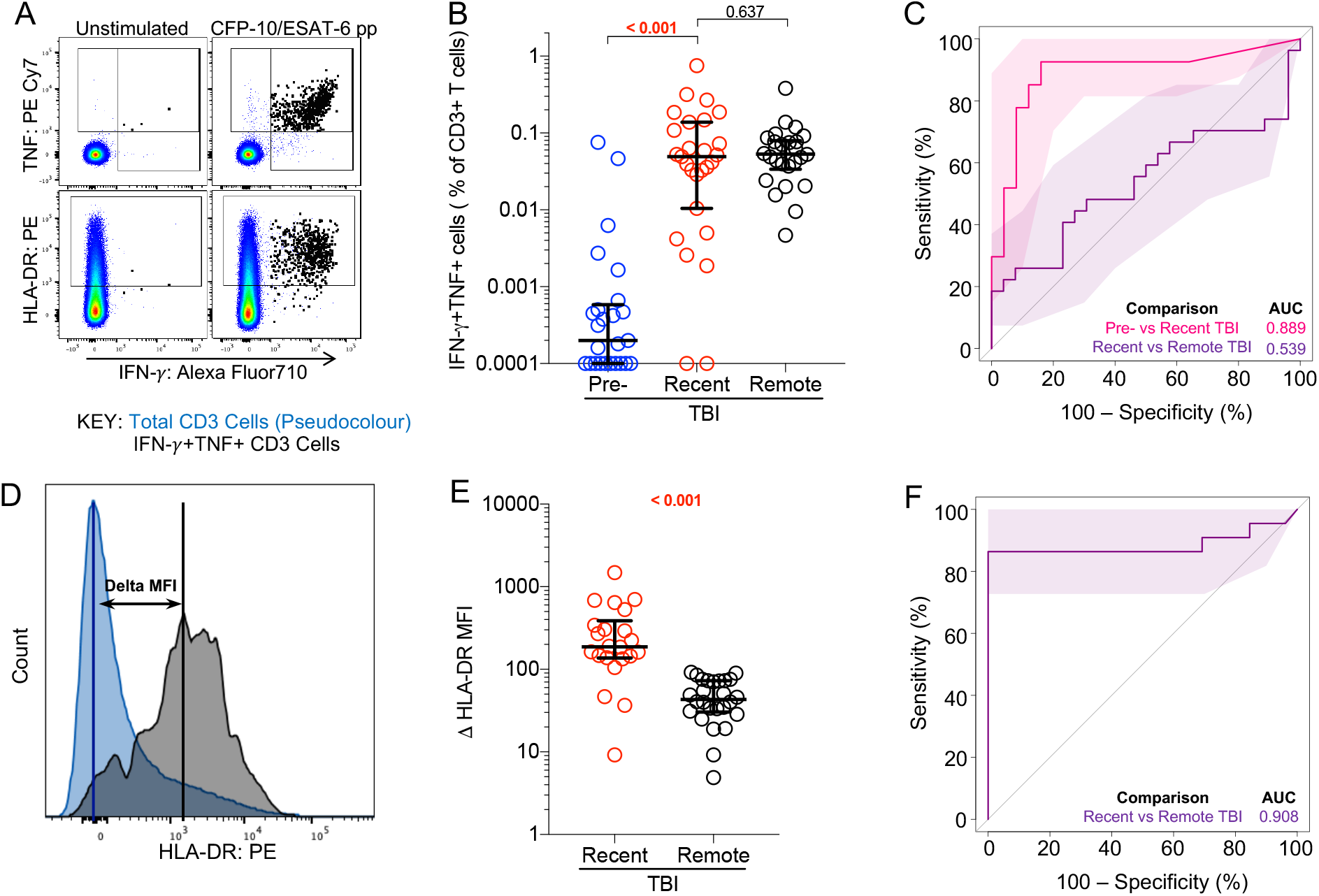
Recent TBI is associated with higher T cell activation than remote TBI. (A) Representative flow cytometry plots depicting IFN-γ, TNF and HLA-DR expression in CD3+ T cells. IFN-γ+TNF+ and total CD3+ T cells are depicted by black and pseudocolour dots, respectively. (B) Frequencies of background subtracted CFP-10/ESAT-6-specific IFN-γ+TNF+ CD3+ T cells detected before (pre-TBI, blue, n=25) and after (recent TBI, red, n=27) infection, and during remote TBI (black, n=26) in the training cohort. (C) Area under the receiving operating characteristic curve (AUCROC) showing performance of CFP-10/ESAT-6-specific IFN-γ+TNF+ CD3+ T cells to discriminate between pre-TBI and recent TBI, and between recent and remote TBI. (D) Representative flow cytometry histogram overlay of HLA-DR expression levels by IFN-γ+TNF+ CD3+ T cells (black) and total CD4 T cells (blue), and how ΔHLA-DR MFI is calculated. (E) ΔHLA-DR MFI in recent (n=22) and remote (n=26) TBI responders in the training cohort. (F) Performance of ΔHLA-DR MFI to discriminate between recent and remote TBI. P-values were calculated using the Wilcoxon-signed rank for paired (pre-TBI *versus* recent TBI) or Mann-Whitney U test for unpaired (recent *versus* remote TBI) comparisons. Where appropriate we corrected for multiple comparison as described in the methods. P-values highlighted in bold and red are considered significant. Shaded areas in AUCROC plots depict 95% confidence intervals. Values less than 0.0001 were set to 0.0001 to allow display on a logarithmic scale.

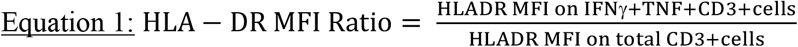

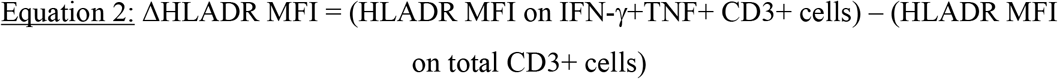

### Statistical analysis

Statistical analyses were performed using R and GraphPad Prism v7. We applied Wilcoxon signed-rank and Mann-Whitney U tests for paired and un-paired analyses, respectively. We considered p values <0.025 as significant, after correcting for multiple comparisons using the Bonferroni method when comparing 3 groups. Functional and activation features identified as significantly different between TBI states were tested for their diagnostic potential using receiver operating characteristic (ROC) curve analysis performed using the pROC package in R (version 1.15.3; https://web.expasy.org/pROC/; 30). Biomarker selection was performed in the training cohort, and tested in a blinded fashion in the test cohort. We selected the ΔHLA-DR MFI threshold for the test cohort based on the best sensitivity between TBI and tuberculosis. This threshold was then applied to compare the performance of the biomarker between recent and remote TBI.

## Results

To identify potential biomarkers of recent TBI, we first evaluated a training cohort of adolescents with recent (n=30) and remote (n=30) TBI (Figure 1A, Supplementary Table 1), and compared T cell polyfunctional, memory and activation profiles between the two TBI states. Out of all the immune features we measured, T cell activation was the best candidate biomarker that distinguished between remote and recent TBI (Mpande et al., manuscript in preparation). With a simplified assay, we then tested the T cell activation biomarker in a blinded test cohort of adolescents with recent (n=25) and remote (n=25) TBI, and adults with tuberculosis disease (n=25) (Figure 1A-B). The T cell activation biomarker was also evaluated with a different method in a cohort of adolescent progressors and non-progressors (Figure 1C). In the training cohort, we measured expression of HLA-DR, IFN-γ and TNF in live CD3+ T cells upon PBMC stimulation with CFP-10/ESAT-6 (Figure 2A, Supplementary Figures 1A and 2A). IFN-γ levels measured by QFT and flow cytometry were strongly correlated (Supplementary Figure 2B). Activation was measured on cytokine-producing T cells as HLA-DR MFI, thus contribution of non-specific signal from the unstimulated control cannot be subtracted. We defined antigen-specific T cells as IFN-γ+TNF+CD3+ cells, based on previous observations that activation measured on CFP-10/ESAT-6-specific IFN-γ+TNF+ could yield better diagnostic accuracy than IFN-γ+ T cells (19). The IFN-γ+TNF+ subset was also associated with lower responses than IFN-γ+ CD3 T cells in unstimulated controls (Supplementary Figure 2A).

Acquisition of TBI (i.e. QFT conversion) was associated with an increase of CFP-10/ESAT-6-specific IFN-γ+TNF+ and IFN-γ+ CD3 T cells, which were detectable at comparable frequencies in recent and remote TBI (Figure 2B, Supplementary Figure 2B). Similar to the QFT assay, frequencies of CFP-10/ESAT-6-specific IFN-γ+TNF+ CD3 T cells allowed discrimination between pre-TBI (i.e. QFT-) and recent TBI (i.e. QFT+) with an area under the ROC curve (AUCROC) of 0.89 [95% confidence interval (CI): 0.79-0.99], but not recent and remote TBI (AUCROC 0.54, 95% CI: 0.38-0.70, Figure 2C). Results for IFN-γ+ CD3 T cells were similar (Supplementary Figure 3C-D).

Analysis of T cell activation in CFP-10/ESAT-6-stimulated samples was restricted to samples that passed our responder criteria (see methods). ΔHLA-DR MFI (Figure 2E), proportions of HLA-DR+ IFN-γ+TNF+ CD3+ T cells (Supplementary Figure 3A) and HLA-DR MFI ratio (Supplementary Figure 3B) were significantly higher in recent TBI compared remote TBI. All biomarkers yielded promising discriminatory potential with AUCROCs of 0.91 (95% CI: 0.81-1.00) for ΔHLA-DR MFI (Figure 2F), 0.93 (95% CI: 0.84-1.00) for % HLA-DR and 0.91 (95% CI: 0.80-1.00) for HLA-DR MFI ratio (Supplementary Figure 3D). We also evaluated the only previously published candidate biomarker for recent TBI, namely proportions of TNF only-expressing effector (CD45RA-CCR7-) CD4+ T cells (26), and found similar levels in recent and remote TBI, with poor diagnostic performance (AUCROC of 0.61; 95% CI: 0.42-0.80; Supplementary Figure 3C-D). We selected ΔHLA-DR MFI for blinded confirmation in the test cohort because it does not require selection of a discrete HLA-DR-positive cell subset using a threshold or “gate”, thus reducing operator bias, and also avoids the issue of negative MFI values that can result from flow cytometric compensation, which would affect calculation of the ratio-based biomarker.

Next, we evaluated performance of the ΔHLA-DR MFI biomarker to distinguish between recent and remote TBI, and between TBI and tuberculosis in the test cohorts. We developed a simplified version of the PBMC-ICS assay and an algorithm to provide a framework for interpreting test results and determine potential TBI states (Table 1). TBI was defined as a positive CFP-10/ESAT-6-specific IFN-γ+TNF+ CD3+ T cell response. In those with TBI, individuals were then classified into recent TBI, remote TBI or those with tuberculosis based on their ΔHLA-DR MFI.

**Table 1:**
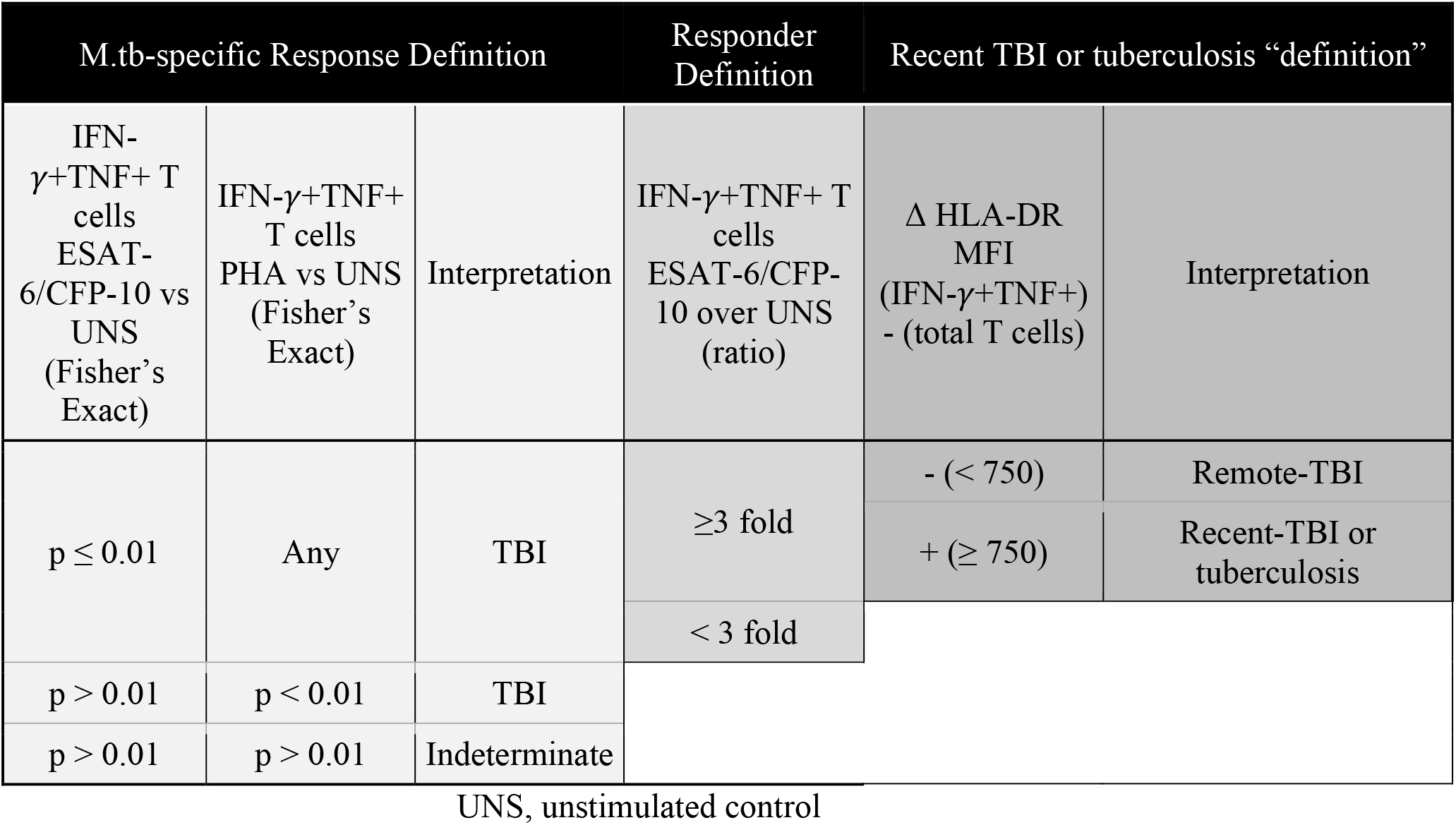
Interpretation of the T cell activation biomarker test results.

CFP-10/ESAT-6-specific IFN-γ+TNF+ CD3+ T cells induced by recent TBI in the test cohorts were detected at similar frequencies to those with remote TBI or with tuberculosis (Figure 3A). Importantly, we were able to discriminate M.tb-uninfected (QFT-) individuals from those with recent or remote TBI (QFT+) with an AUCROC of 0.94 (95% CI: 0.87-1.00; Figure 3B), suggesting that this simple flow cytometry-based assay yields equivalent diagnostic information to QFT.

**Figure 3:**
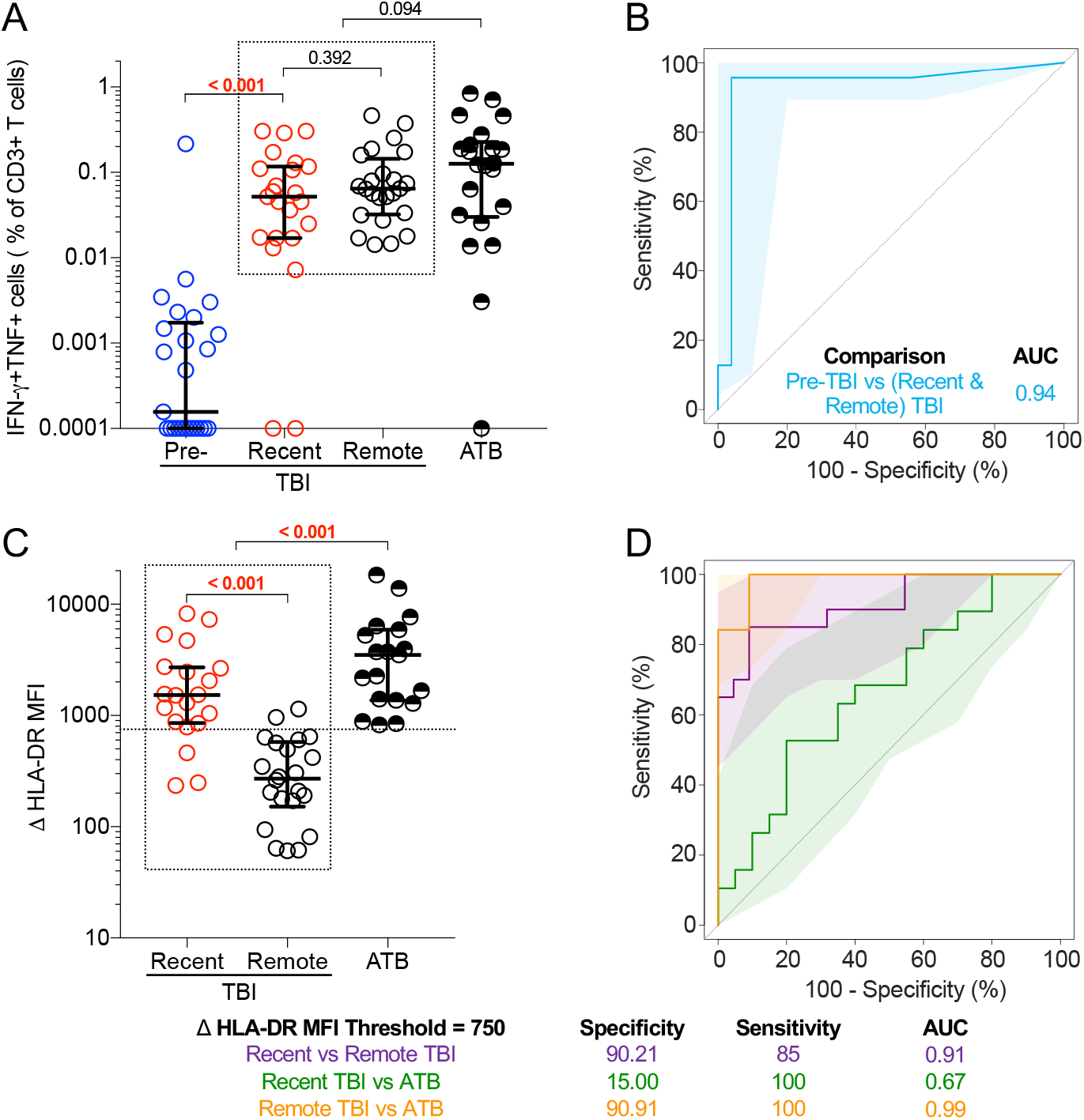
T cell biomarkers can distinguish between recent and remote TBI, as well as between TBI and tuberculosis. (A) Frequencies of background subtracted CFP-10/ESAT-6-specific IFN-γ+TNF+ CD3+ T cells detected before (pre-TBI, blue, n=25) and after (recent TBI, red, n=23) TBI, during remote TBI (black open symbols, n=24), and in tuberculosis patients (ATB, black half-filled symbols, n=22) in the test cohort. (B) ROC curve analysis illustrating the performance of CFP-10/ESAT-6-specific IFN-γ+TNF+ CD3+ T cells to distinguish between samples taken before TBI (pre-TBI) and those taken after TBI (recent or remote TBI combined). (C) ΔHLA-DR MFI in responders with recent TBI (n=20), remote TBI (n=22) or tuberculosis (ATB, n=19). (D) ROC curve analysis depicting the performance of ΔHLA-DR MFI to discriminate between recent and remote TBI, between recent TBI and tuberculosis (ATB) and between remote TBI and tuberculosis. P-values were calculated using the Wilcoxon-signed rank for paired (pre-TBI *versus* recent TBI) or the Mann-Whitney U test for unpaired (all other) comparisons. Where appropriate we corrected for multiple comparisons as described in the methods. P-values highlighted in bold and red are considered significant. Shaded areas in AUCROC plots depict 95% confidence intervals. Values less than 0.0001 were set to 0.0001 to allow display on a logarithmic scale.

ΔHLA-DR MFI was significantly higher in recent TBI compared to remote TBI (Figure 3C), and in tuberculosis patients compared to individuals with TBI (Figure 3C). Using a ΔHLA-DR MFI cut-off of 750, we were able to discriminate between recent and remote TBI with a specificity, sensitivity and AUCROC of 90%, 85% and 0.91 (95% CI: 0.83-1.00), respectively. We also observed excellent discrimination between remote TBI and tuberculosis, with high specificity (91%), sensitivity (100%) and AUCROC (0.99, 95% CI: 0.96-1.00). ΔHLA-DR MFI could not accurately discriminate between recent TBI and tuberculosis (AUC=0.67, 95% CI: 0.50-0.84; Figure 3C-D). Supplementary Table 6 summarises the outcomes of the diagnostic algorithm described in Table 1 when applied to the test cohort.

Our group previously reported a 6-gene blood transcriptomic signature, RISK6, that can identify individuals with TBI who are at risk of progressing to tuberculosis (31). We therefore tested the ability of the ΔHLA-DR MFI biomarker to distinguish between progressors (n=34) and non-progressors (n=34) in a longitudinal cohort of healthy, TBI adolescents (Figure 1C). Levels of ΔHLA-DR MFI were significantly higher in progressors up to 2 years before tuberculosis diagnosis compared to non-progressors (AUCROC of 0.75; 95% CI: 0.63-0.87; Figure 4). These results were consistent with the performance of RISK6 and suggest that the ΔHLA-DR MFI biomarker may also be useful to identify individuals at risk of disease progression.

**Figure 4:**
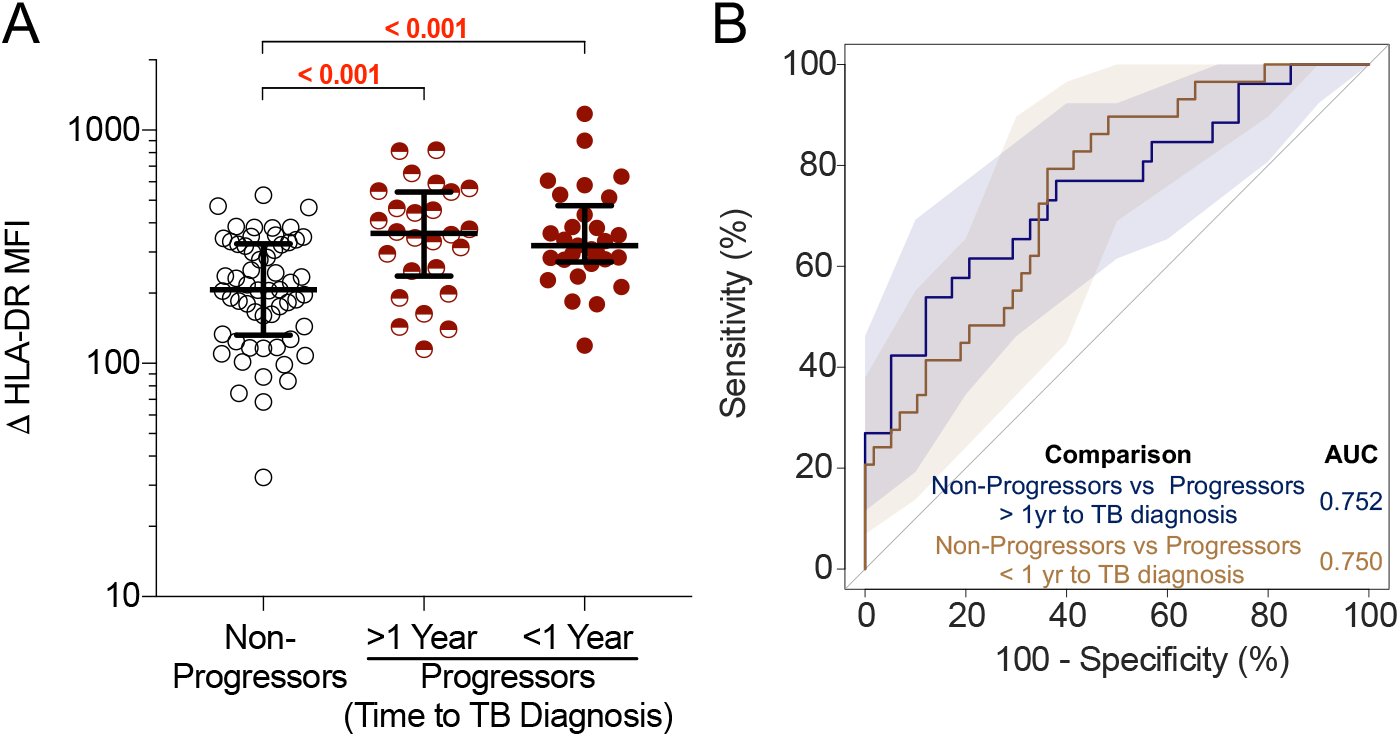
Mycobacteria-specific T cells in tuberculosis progressors are more activated than those in non-progressors. (A) ΔHLA-DR MFI on Mtb lysate-responsive T cells co-expressing *IFNG* and *TNF* mRNA transcripts in non-progressors (n=58 longitudinal data points from 34 participants) and progressors [in samples collected >1 year before tuberculosis diagnosis (> 1 Year, n=26 longitudinal data points from 19 participants) or in samples collected within 1 year of tuberculosis diagnosis (< 1 Year, n=29 longitudinal data points from 22 participants)]. P-values were computed by Mann–Whitney U test and were corrected for 2 comparisons as described in the methods. (B) ROC curve (AUC) depicting the performance of ΔHLA-DR MFI to discriminate between non-progressors and progressors in samples collected >1 year before tuberculosis or within 1 year of tuberculosis diagnosis.

## Discussion

We describe a simple biomarker, ΔHLA-DR MFI, that measures M.tb-specific T cell frequencies and their activation levels, and can distinguish M.tb uninfected individuals from those with TBI, and among the latter can identify those with recent TBI, disease progression and active tuberculosis. This is a significant advance over current M.tb immunodiagnostics (TST and IGRA), which measure immune responses and are unable to distinguish between TBI and tuberculosis, between recent and remote TBI or between TBI and tuberculosis progressors. Consistent with this limitation, we demonstrated that measuring M.tb-specific T cell functions by flow cytometry can distinguish between IGRA- and IGRA+ individuals, but not between TBI and tuberculosis, nor recent and remote TBI. Since individuals with recent TBI and high risk for disease progression would benefit from targeted TPT, this biomarker offers an opportunity to identify a group of asymptomatic individuals at particularly high risk of tuberculosis, while those with remote TBI would derive less benefit from TPT. This is of particular importance in high incidence settings, where a large proportion of the population is already M.tb infected and identification of individuals at high risk of TB progression is challenging. Our findings build on previous studies that have shown that measurement of M.tb-specific T cell activation by flow cytometry has promising diagnostic potential to distinguish TBI from tuberculosis disease, regardless of HIV status, as well as to monitor treatment response (16-19).

The immunological hypothesis that underpins our observations is that T cell activation reflects M.tb antigen load *in vivo*, and that these levels peak during primary infection and then drop in most people with remote infection, due to control of bacterial replication. However, in people who progress towards active tuberculosis, M.tb antigen load rises again to reach high levels in those with clinical tuberculosis. Our measurements of T cell activation during the different stages of TBI provide evidence that supports this hypothesis. Further, our findings suggest that remote TBI is associated with low and stable levels of M.tb-specific T cell activation, consistent with control or perhaps even clearance of M.tb. This supports the hypothesis that tuberculosis has a short incubation period (<2 years) and that M.tb-specific T cell responses detected with IGRAs in most individuals with remote TBI represent immunological memory, and not ongoing M.tb replication (13). Therefore, our biomarker has utility to distinguish the majority of individuals with remote TBI and low risk of tuberculosis progression, who would be spared TPT, from the minority who harbour replicating M.tb, are at high risk of progression, and would benefit from further clinical investigation and TPT.

The strengths of our study include unique and well-characterized clinical cohorts, blinded verification of our findings in a test cohort and translation of biological observations made with complex technology to a simple test that can be used for further validation.

Limitations of our study include the small and retrospective case-control study design, and that all adolescent participants were selected from the same epidemiological study (10). We were thus unable to formally validate our ΔHLA-DR MFI biomarker because the test, training and progressor cohorts were not completely independent. The control group of newly diagnosed tuberculosis included adults aged 21-42 years, therefore we cannot exclude age-related differences. Further, experimental protocols used to detect T cell activation were different between cohorts. While the consistency of results points towards robustness of the biomarker, this experimental setup prevented us from defining a positivity cut-off in the training cohort, and formally validating it in the test cohorts. Additionally, samples from the progressor cohort were stimulated with M.tb lysate, which contains antigens cross-reactive with other mycobacteria, and included sorting of reactive cells prior to measurement of IFN-γ and TNF expression. These factors could have contributed to lower discriminatory power between progressors and non-progressors.

The candidate biomarker proposed by Halliday and colleagues (TNF only-expressing TEFF cells; 26) did not discriminate between recent and remote TBI in our study. Compared to the original biomarker description, we used a different flow cytometry panel that lacked CD127, and therefore the cell subset definition was not identical.

Finally, experimental protocols described here were performed on cryopreserved PBMC, a sample type that may not be ideal for clinical purposes, but was the only one available for this retrospective study. Although this is not a point-of-care assay, we have shown that it can be adapted to a whole blood assay (such as QFT) with a simple 4-colour flow cytometry panel as readout (19), using basic technology that is widespread even in resource limited settings and is suitable for automated analysis. It will be necessary to validate this simplified assay in large field studies to estimate its true performance characteristics. Preferred performance characteristics of a test that can differentiate between recent and remote TBI are currently not defined and would need to be developed to provide benchmarks for test performance. A validation is currently underway in a prospective paediatric cohort of house-hold contacts of tuberculosis patients.

In summary, we describe a blood-based T cell biomarker, ΔHLA-DR MFI, measured with a simple flow cytometry assay, that can be used to identify individuals with recent TBI and those with tuberculosis progression, as well as clinical disease. Upon further validation, the ΔHLA-DR MFI biomarker has the potential to stratify individuals along the M.tb spectrum and could be considered as a rule-in test, to identify individuals at high risk of tuberculosis for further evaluation. If interpreted in association with clinical features and microbiological tests, this biomarker could identify individuals with tuberculosis who would benefit from standard treatment, those at high risk who should receive TPT, and those who would not currently benefit from TPT but may be re-tested in future.

## Data Availability

Data will be available once the pre-print is published in a peer-reviewed journal

## Contributors

MMD, MR, MH, TJS and EN designed the study and raised funding; CAMM, MM, BM, TDR, CS, NB, HH and GO generated the data; CAMM, MM, VR, TL and HH analysed the data; CAMM, MM, VR, MMD, MR, MH, TJS and EN interpreted the results; CAMM, MM, VR, TJS and EN wrote the manuscript. All authors revised and approved the manuscript and are accountable for the work.

## Funding Statement

US National Institutes of Health (R21AI127121), Bill and Melinda Gates Foundation (BMGF; OPP1066265, OPP1113682) and the South African Medical Research Council funded the study. The South African National Research Foundation and Carnegie Corporation funded scholarships to CAMM and MM, respectively. The ACS study was supported by Aeras and BMGF (GC 6-74: grant 37772; GC12: grant 37885) for QFT testing. The funding sources has no role in study design, data collection, analysis and interpretation, writing and submission of the manuscript.

## Declarations of interests

EN reports grants to the University of Cape Town from AERAS, Bill & Melinda Gates Foundation and NIH during the conduct of the study. TJS reports grants to the University of Cape Town from AERAS, Bill & Melinda Gates Foundation, NIH and the SAMRC during the conduct of the study.

All other authors declare no competing interests.

## Acknowledgments

We are grateful to the study participants and their families; the Cape Winelands East district communities, the Department of Education and the Department of Health; the SATVI clinical and laboratory teams; Thomas Hawn and Jason Andrews for valuable input on the study design.

## Supplementary methods

### PBMC stimulation and staining protocols

#### Recent and remote TBI Protocol

##### Stimulation

Cells were thawed, rested for 4 hours and stimulated in R10 media (RPMI, 10% Fetal Bovine Serum, 1% L-glutamine and 1% penicillin-streptomycin) containing anti-CD107a (training cohort only) with either no antigen (unstimulated), CFP-10/ESAT-6 peptide pool (15 mer peptides overlapping by ten amino-acids, 1μg/mL, GenScript Biotechnology), Staphylococcal enterotoxin B (SEB, 1μg/mL, Sigma Aldricht, training cohort) or phytohemagglutinin (5μg/mL, Remel, test cohort). Cells were incubated for 3 hours at 37°C with 5% CO2, after which brefeldin A (BFA, 5μg/mL, Sigma Aldrich) and monensin (2.5μg/mL, Sigma Aldrich) were added and incubated for a further 15 hours (training cohort). BFA was added at the onset of the 18 hour stimulation in the test cohort. *Staining* (Supplementary Table 2 and 3): Cells were treated with PBS 2% EDTA, washed and stained with anti-CCR7 and anti-CXCR3 for 30 minutes at 37°C (training cohort only). Staining for viability and surface markers was performed for 30 minutes at room temperature. Cells were washed, fixed and permeabilised (CytoFix/CytoPerm, BD Biosciences) prior to intra-cellular staining of functional markers for 30 minutes at room temperature. Cells were washed and fixed (PBS 1% paraformaldehyde) prior to acquisition on a LSRII flow cytometer (BD Biosciences; Supplementary Figure 1A).

#### Progressor and Non-progressor Protocol

Cryopreserved PBMC from progressors and non-progressors were thawed, rested for 6 hours, and stimulated in R10 for 12 hours with M.tb lysate (final concentration 10µg/mL, BEI Resources: NR-14822) in the presence of anti-CD49d antibody (final concentration 1µg/mL), and anti-CD154-PE (final concentration 10µL/mL). Following stimulation cells were washed and stained with LIVE/DEAD Fixable Aqua Stain for 30 minutes at 4°C. Next, cells were washed and stained with antibodies (Supplementary Table 4) for 60 minutes at room temperature. Activated (i.e. CD69+CD137+ and/or CD69+CD154+) single T cells were index-sorted (BD FACS Aria-II; Supplementary Figure 1B) into 96 well plates containing One-Step RT-PCR buffer (Qiagen) and a panel of primers specific for 20 mRNA transcripts (Supplementary Table 5). Transcript-specific amplification was performed and further amplified in a nested PCR before sequencing on a MiSeq (Illumina) instrument (1,2).

### Biomarker Definition

#### Candidate biomarker

Halliday and colleagues showed that the proportion of TNF+IL-2-IFN-γ-CD4 T cells with an effector memory phenotype (TE), defined as CD45RA-CCR7-CD127-, detected by flow cytometry after stimulation with M.tb purified protein derivative, could distinguish recent from remote TBI (3). Since we did not include CD127 in our flow cytometry panel, we defined TNF+IL-2-IFN-γ-TE memory CD4 T cells as CD45RA-CCR7-. We considered those individuals with frequencies of antigen-specific CD4 T cells expressing any combination of Th1 cytokines (IFN-γ±TNF±IL-2±) that were significantly higher [false discovery rate ≤0.01, calculated by MIMOSA in R, version 1.21.0; https://github.com/RGLab/MIMOSA (4); and at least 3 fold increase] than background as responders. Non-responders were excluded from the analysis.

---

**Supplementary Figure 1:**
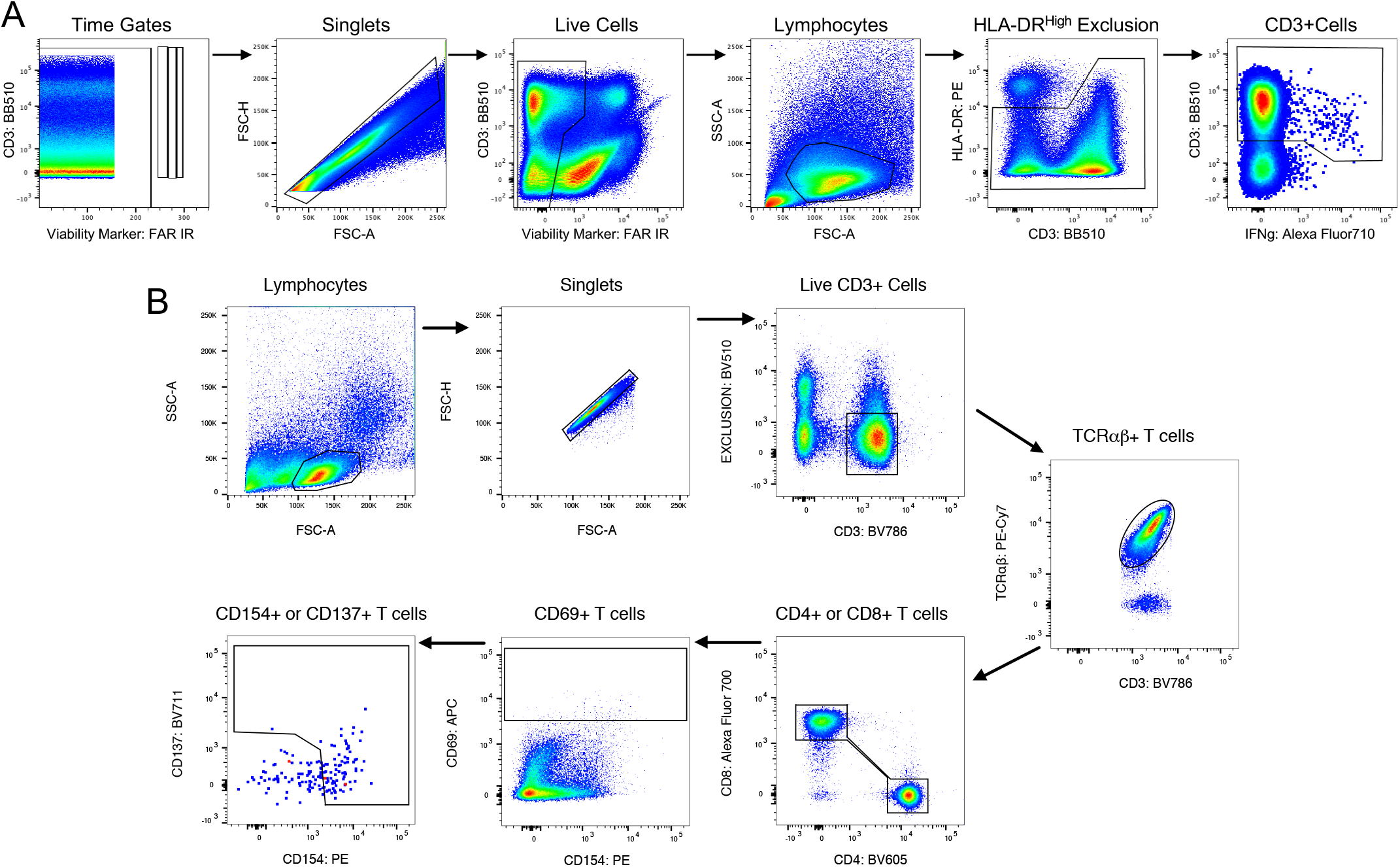
Gating strategy used to identify M.tb-responsive T cells. Gating strategies used for training and test cohorts were similar. We first gated on total cells based on consistency of fluorescence over acquisition time (time gate), followed by exclusion of cell doublets (singlet gate), dead cells and antibody aggregates. We then gated on total lymphocytes, excluded CD3-cells that expressed high levels of HLA-DR, which we assumed were B cells, and then gated on total CD3+ cells. Functional markers (IFN-γ and TNF) and activation marker HLA-DR were then selected among total CD3+ cells as indicated in Figure 2A. (B) Gating strategy used to identify M.tb-responsive T cells following stimulation with Mtb lysate in the progressor and non-progressor cohort. Live, Mtb-responsive αβ T cells were defined as CD69+ cells co-expressing CD154 and/or CD137. M.tb-responsive T cells were index sorted into 96-well plates for single cell gene expression profiling.

**Supplementary Figure 2:**
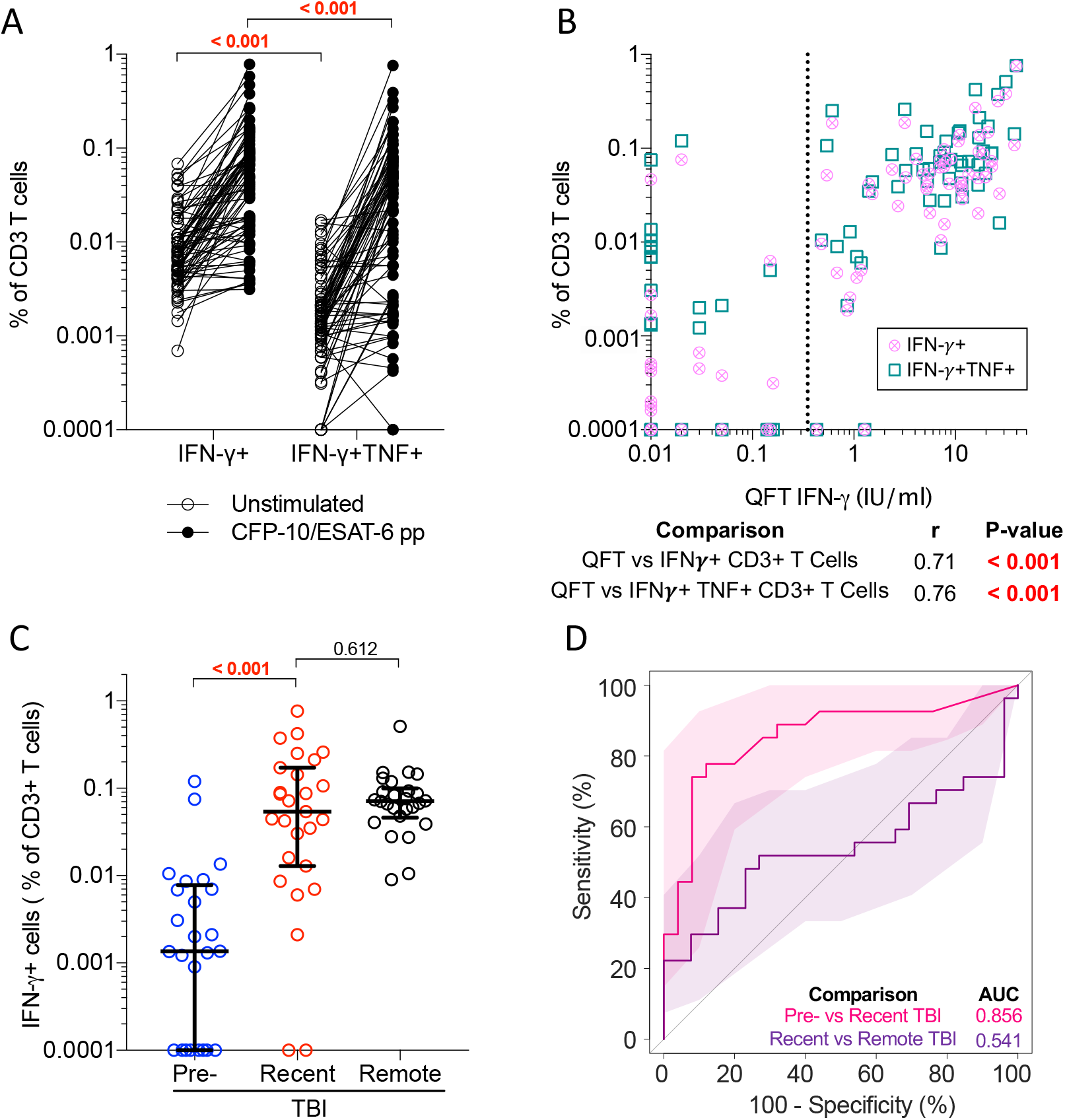
Functional CD3 T cell responses cannot distinguish between recent and remote TBI. (A) Frequencies of unstimulated (open circles, n=78) and CFP-10/ESAT-6-stimulated (black circles, n=78) IFN-γ+ and IFN-γ+TNF+ CD3+ T cells detected across all samples in the training cohort. (B) Frequencies of IFN-γ+ (circles) and IFN-γ+TNF+ (squares) CD3+ T cells detected in all individuals from the training cohort (n=78) by PBMC-ICS were compared to IFN-γ levels (IU/mL) measured by QFT in fresh whole blood. Dotted line at 0.35IU/mL represents the QFT cut-off. r represents the Spearman’s correlation coefficient. (C) Frequencies of background subtracted CFP-10/ESAT-6-specific IFN-γ+ CD3+ T cells detected before (pre-TBI, blue, n=25) and after TBI (recent TBI, red, n=27) and during remote TBI (n=26, black). (C) ROC curve (AUC) depicting the performance of CFP-10/ESAT-6-specific IFN-γ+ CD3 T cells to discriminate between pre-TBI and recent TBI and between recent and remote TBI. P-values were calculated using the Wilcoxon-signed rank test for paired and the Mann-Whitney U test for unpaired comparisons. Where appropriate, p-values were corrected for multiple comparison as described in the methods. P-values highlighted in bold and red were considered significant. Shaded areas depict 95% confidence intervals. Values less than 0.0001 were set to 0.0001 to allow display on a logarithmic scale.

**Supplementary Figure 3:**
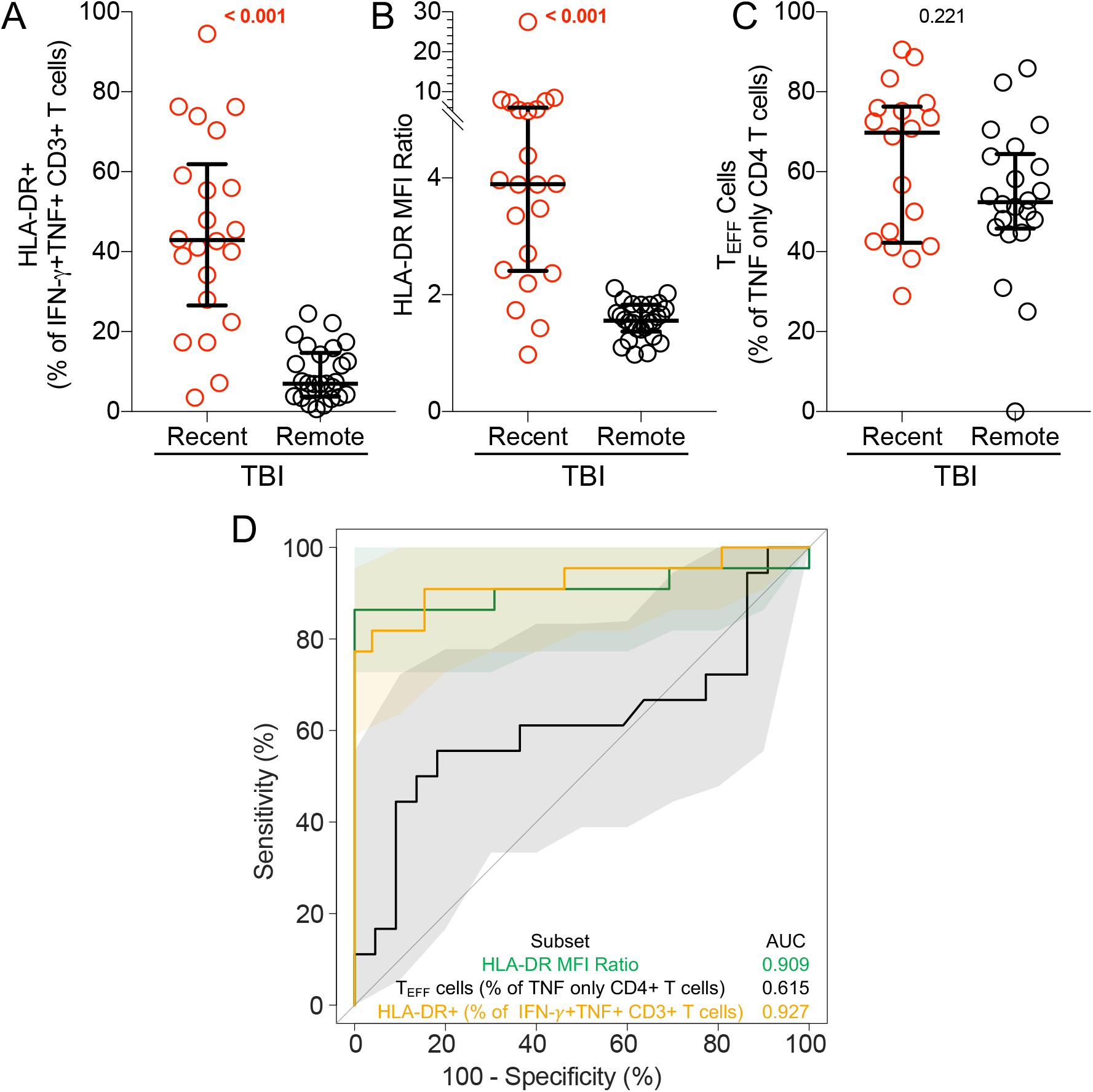
T cell activation but not TNF only TEFF CD4+ T cells can distinguish between recent and remote TBI. Graphs depict (A) HLA-DR+ as a proportion of IFN-γ+TNF+ CD3+ T cells and (B) HLA-DR MFI ratio in recent (n=22) and remote (n=26) TBI. (C) depicts TE (CD45RA-CCR7-) cells as a proportion of TNF+ IFN-γ-IL-2- (TNF only) CD4+ T cells in adolescents with recent (n=18) and remote (n=22) TBI. (D) ROC curve with 95% CI depicting the performance of each biomarker illustrated in A-C to discriminate between pre-TBI and recent TBI and between recent and remote TBI

**Supplementary Table 1:** Demographics.

|  | Training Cohort |  | Test Cohort |  |  | TB Progressor Cohort |  |
| --- | --- | --- | --- | --- | --- | --- | --- |
|  | Remote TBI | Recent TBI | Remote TBI | Recent TBI | aTB | Progressors | Non-progressors |
| **n** | 30 | 30 | 25 | 25 | 25 | 39 | 39 |
| **Gender (Female)** | 21 | 19 | 12 | 9 | 3 | 14 | 28 |
| **Age in years (IQR)** | 15 (14-16) | 15 (14-16) | 15 (14-16) | 15 (14-16) | 32 (21-42) | 16 (15-17) | 16 (14-17) |
| **Ethnicity: Coloured** | 26 | 29 | 23 | 23 | 14 | 36 | 37 |
| **Ethnicity: Black** | 4 | 1 | 2 | 1 | 11 | 3 | 2 |
| **Ethnicity: Indian** | 0 | 0 | 0 | 1 | 0 | 0 | 0 |

**Supplementary Table 2:** Training Cohort Flow Cytometry Panel.

|  | Role | Fluorochrome | Clone | Manufacturer | Cat. Number |
| --- | --- | --- | --- | --- | --- |
| CD3 | Lineage | BV650 | UCHT1 | BD Biosciences | 563852 |
| CD4 |  | BV785 | OKT4 | Biolegend | 317442 |
| CD8 |  | BV711 | RPA-T8 | BD Biosciences | 301044 |
| CCR7 | T cell differentiation | PE | 150503 | BD Biosciences | 560765 |
| CD27 |  | BV510 | L128 | BD Biosciences | 563092 |
| CD45RA |  | BV570 | HI100 | eBioscience | 304132 |
| KLRG-1 |  | PercP-eFlour710 | 13F12F2 | eBioscience | 46948842 |
| CXCR3 | T<sub>SCM</sub>; Homing | PE-Cy5 | 1C6/CXCR3 | BD Biosciences | 551128 |
| HLA-DR | Activation | FITC | L243 | BD Biosciences | 307604 |
| CD107 | Function | PE-CF594 | H4A3 | BD Biosciences | 562628 |
| CD154 (CD40L) |  | BV421 | TRAP-1 | BD Biosciences | 563886 |
| IFN-γ | | Alexa Fluor 700 | B27 | BD Biosciences | 557995 |
| IL-2 |  | APC | Rat | BD Biosciences | 554567 |
| TNF |  | PE-Cy7 | Mab11 | BD Biosciences | 25734982 |
| Live/Dead | Viability | Near IR (APC-H7) | N/A | Life Technologies | L34976 |

Markers highlighted in grey were not used for the purpose of the analysis reported.

**Supplementary Table 3:** Test Cohort Flow Cytometry Panel.

| Marker | Clone | Fluorochrome | Clone | Manufacturer | Cat. Number |
| --- | --- | --- | --- | --- | --- |
| CD3 | Lineage | Brilliant Blue 515 | UCHT1 | BD | 564465 |
| HLA-DR | Activation | PE | L243 | BD | 347401 |
| IFN-γ | Function | APC | B27 | BD | 554702 |
| TNF | Function | PE-Cy7 | Mab11 | eBioscience | 25734982 |
| Live/Dead | Viability | Near IR (APC-H7) | N/A | Life Technologies | L34976 |

**Supplementary Table 4:** TB-Progressor Cohort Flow Cytometry Panel.

| Marker | Clone | Fluorochrome | Clone | Manufacturer | Cat. Number |
| --- | --- | --- | --- | --- | --- |
| CD3 | Lineage | BV786 | SK7 | BD Biosciences | 563800 |
| CD4 |  | BV605 | RPA-T4 | Biolegend | 300556 |
| CD8 |  | Alexa Fluor 700 | RPA-T8 | BD Biosciences | 561453 |
| TCR$\alpha\beta$ | | PE-Cy7 | IP26 | Biolegend | 306720 |
| CD69 | Antigen Specificity | APC | L78 | BD Biosciences | 340560 |
| CD137 |  | BV711 | 4B4-1 | BD Biosciences | 740798 |
| CD154 (CD40L) |  | PE | TRAP1 | BD Biosciences | 555700 |
| CD26 | Phenotype | FITC | BA5b | Biolegend | 302704 |
| HLA-DR | Activation | BV421 | L243 | Biolegend | 307636 |
| CD14 | Exclusion | BV510 | M5E2 | Biolegend | 301842 |
| CD19 |  | BV510 | HIB19 | Biolegend | 302242 |
| Live/Dead | Viability | BV510 | N/A | Invitrogen | L34957 |

Markers highlighted in grey were not used for the purpose of the analysis reported.

**Supplementary Table 5:** mRNA transcripts targeted by single cell phenotyping primers.

|  |  |  |  |  |
| --- | --- | --- | --- | --- |
| *BCL6* | *GZMB* | *IL13* | *IL4* | *RUNX3* |
| *EOMES* | *IFNG* | *IL17A* | *PERF* | *TBET* |
| *FOXP3* | *IL10* | *IL2* | *RORC* | *TGFB* |
| *GATA3* | *IL12A* | *IL21* | *RUNX1* | *TNF* |

**Supplementary Table 6:**
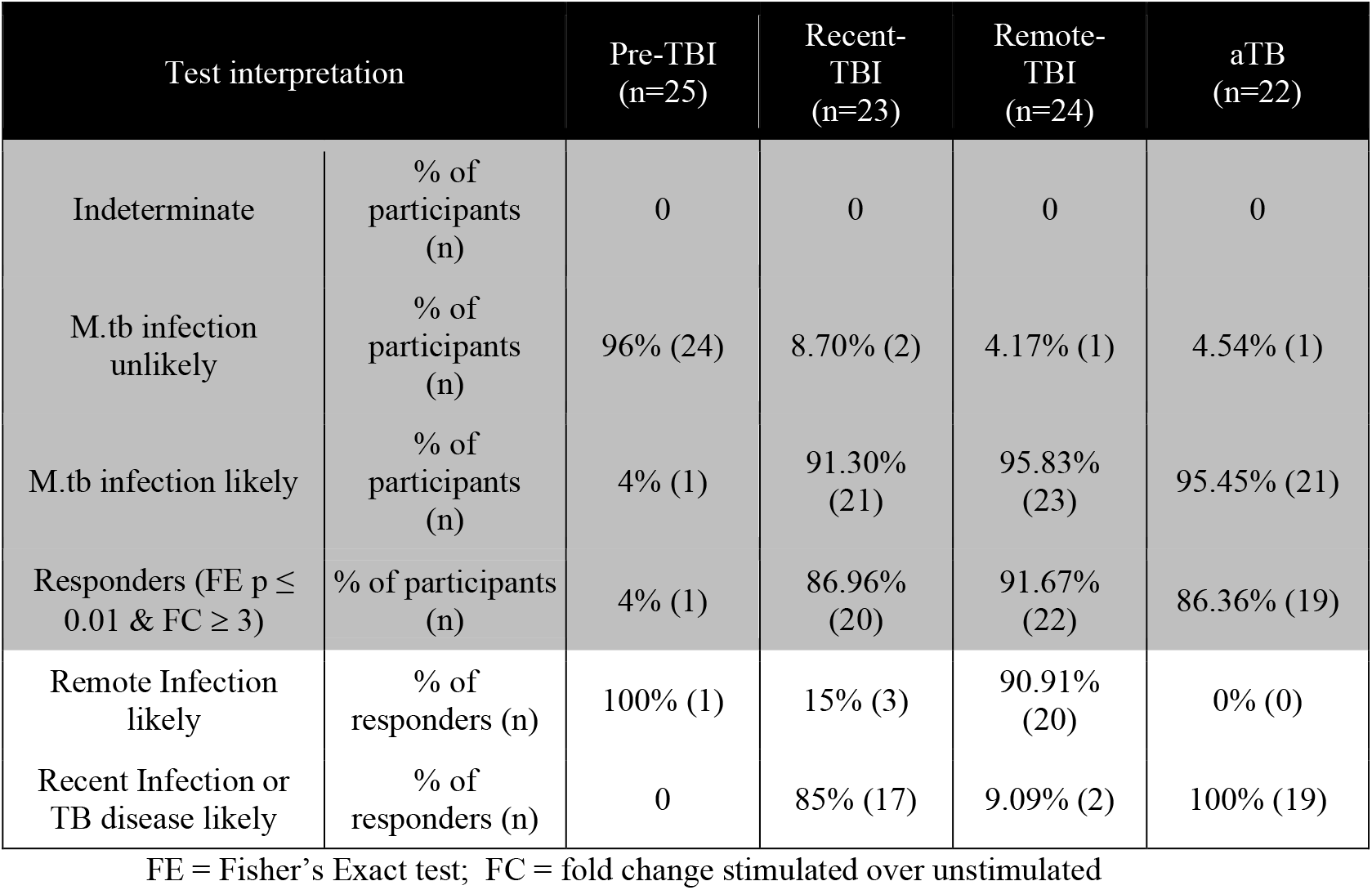
ΔHLA-DR MFI biomarker results.

| Test interpretation |  | Pre-TBI<br>(n=25) | Recent-TBI<br>(n=23) | Remote-TBI<br>(n=24) | aTB<br>(n=22) |
| --- | --- | --- | --- | --- | --- |
| Indeterminate | % of participants (n) | 0 | 0 | 0 | 0 |
| M.tb infection unlikely | % of participants (n) | 96% (24) | 8.70% (2) | 4.17% (1) | 4.54% (1) |
| M.tb infection likely | % of participants (n) | 4% (1) | 91.30% (21) | 95.83% (23) | 95.45% (21) |
| Responders (FE $p \leq 0.01$ & FC $\geq 3$) | % of participants (n) | 4% (1) | 86.96% (20) | 91.67% (22) | 86.36% (19) |
| Remote Infection likely | % of responders (n) | 100% (1) | 15% (3) | 90.91% (20) | 0% (0) |
| Recent Infection or TB disease likely | % of responders (n) | 0 | 85% (17) | 9.09% (2) | 100% (19) |

FE = Fisher's Exact test; FC = fold change stimulated over unstimulated

## Notes

### Competing Interest Statement

The authors have declared no competing interest.

## References

1 World Health Organisation. Global tuberculosis report 2019. Licence: CC BY-NC-SA 3.0 IGO. https://www.who.int/tb/publications/global_report/en/ (xAccessed June 18 2020)

2 World Health Organisation. WHO consolidated guidelines on tuberculosis: tuberculosis preventive treatment. 2020. Licence: CC BY-NC-SA 3.0 IGO https://www.who.int/publications-detail/who-consolidated-guidelines-on-tuberculosis-module-1-prevention-tuberculosis-preventive-treatment (Accessed June 18 2020)

3 Erkens CGM, Slump E, Verhagen M, Schimmel H, Cobelens F, Hof S van den. Risk of developing tuberculosis disease among persons diagnosed with latent tuberculosis infection in the Netherlands. Eur Respir J 2016; 48: 1420–8.

4 Reichler MR, Khan A, Sterling TR, et al. Risk Factors for Tuberculosis and Effect of Preventive Therapy Among Close Contacts of Persons with Infectious Tuberculosis. Clin Infect Dis 2019; 70: 1562–72.

5 Cohen A, Mathiasen VD, Schön T, Wejse C. The global prevalence of latent tuberculosis: a systematic review and meta-analysis. Eur Respir J 2019; 54: 1900655.

6 Mathema B, Andrews JR, Cohen T, et al. Drivers of Tuberculosis Transmission. J Infect Dis 2017; 216: S644–53.

7 Churchyard G, Kim P, Shah NS, et al. What We Know About Tuberculosis Transmission: An Overview. J Infect Dis 2017; 216: S629–35.

8 Wiker HG, Mustafa T, Bjune GA, Harboe M. Evidence for waning of latency in a cohort study of tuberculosis. BMC Infect Dis 2010; 10: 37.

9 Behr MA, Edelstein PH, Ramakrishnan L. Revisiting the timetable of tuberculosis. BMJ 2018; 362: k2738.

10 Machingaidze S, Verver S, Mulenga H, et al. Predictive Value of Recent QuantiFERON Conversion for Tuberculosis Disease in Adolescents. Am J Resp Crit Care 2012; 186: 1051–1056.

11 Andrews JR, Noubary F, Walensky RP, Cerda R, Losina E, Horsburgh CR. Risk of Progression to Active Tuberculosis Following Reinfection With Mycobacterium tuberculosis. Clin Infect Dis 2012; 54: 784–791.

12 Cadena AM, Hopkins FF, Maiello P, et al. Concurrent infection with Mycobacterium tuberculosis confers robust protection against secondary infection in macaques. Plos Pathog 2018; 14: e1007305.

13 Behr MA, Edelstein PH, Ramakrishnan L. Is Mycobacterium tuberculosis infection life long? BMJ 2019; 367: 5770.

14 World Health Organisation. FIND. Consensus Meeting Report: Development of a Target Product Profile (TPP) and a framework for evaluation for a test for predicting progression from tuberculosis infection to active disease. 2017 (WHO/HTM/TB/2017.18). https://apps.who.int/iris/handle/10665/259176 (Accessed June 18 2020)

15 STOP TB Partnership. Global Plan to End TB: 2018-2022. http://www.stoptb.org/assets/documents/global/plan/GPR_2018-2022_Digital.pdf (Accessed June 18 2020)

16 Adekambi T, Ibegbu CC, Cagle S, et al. Biomarkers on patient T cells diagnose active tuberculosis and monitor treatment response. J Clin Invest 2015; 125: 1827–1838.

17 Wilkinson KA, Oni T, Gideon HP, Goliath R, Wilkinson RJ, Riou C. Activation Profile of Mycobacterium tuberculosis–Specific CD4+ T Cells Reflects Disease Activity Irrespective of HIV Status. Am J Resp Crit Care 2016; 193: 1307–10.

18 Riou C, Berkowitz N, Goliath R, Burgers WA, Wilkinson RJ. Analysis of the Phenotype of Mycobacterium tuberculosis-Specific CD4+ T Cells to Discriminate Latent from Active Tuberculosis in HIV-Uninfected and HIV-Infected Individuals. Front Immunol 2017; 8: 968.

19 Musvosvi M, Duffy D, Filander E, et al. T-cell biomarkers for diagnosis of tuberculosis: candidate evaluation by a simple whole blood assay for clinical translation. Eur Respir J 2018; 51: 1800153.

20 Portevin D, Moukambi F, Clowes P, et al. Assessment of the novel T-cell activation marker–tuberculosis assay for diagnosis of active tuberculosis in children: a prospective proof-of-concept study. Lancet Infect Dis 2014; 14: 931–8.

21 Petruccioli E, Petrone L, Vanini V, et al. Assessment of CD27 expression as a tool for active and latent tuberculosis diagnosis. J Infection 2015; 71: 1–8.

22 Harari A, Rozot V, Enders FB, et al. Dominant TNF-α+ Mycobacterium tuberculosis– specific CD4+ T cell responses discriminate between latent infection and active disease. Nat Med 2011; 17: 372–376.

23 Rozot V, Patrizia A, Vigano S, et al. Combined Use of Mycobacterium tuberculosis– Specific CD4 and CD8 T-Cell Responses Is a Powerful Diagnostic Tool of Active Tuberculosis. Clin Infect Dis 2015; 60: 432–7.

24 Andersen P, Doherty TM, Pai M, Weldingh K. The prognosis of latent tuberculosis: can disease be predicted? Trends Mol Med 2007; 13: 175182.

25 Scriba TJ, Penn-Nicholson A, Shankar S, et al. Sequential inflammatory processes define human progression from M. tuberculosis infection to tuberculosis disease. Plos Pathog 2017; 13: e1006687.

26 Halliday A, Whitworth H, Kottoor SH, et al. Stratification of Latent Mycobacterium tuberculosis Infection by Cellular Immune Profiling. J Infect Dis 2017; 215: 1480–1487.

27 Zak DE, Penn-Nicholson A, Scriba TJ, et al. A blood RNA signature for tuberculosis disease risk: a prospective cohort study. Lancet 2016; 387: 2312–2322.

28 Han A, Glanville J, Hansmann L, Davis MM. Linking T-cell receptor sequence to functional phenotype at the single-cell level. Nat Biotechnol 2014; 32: 684–92.

29 Glanville J, Huang H, Nau A, et al. Identifying specificity groups in the T cell receptor repertoire. Nature 2017; 547: 94–8.

30 Robin X, Turck N, Hainard A, et al. pROC: an open-source package for R and S+ to analyze and compare ROC curves. BMC Bioinformatics 2011; 12: 77.

31 Penn-Nicholson A, Mbandi SK, Thompson E, et al. RISK6, a 6-gene transcriptomic signature of TB disease risk, diagnosis and treatment response. Sci Rep 2020; 10: 8629.

## References

1 Glanville J, Huang H, Nau A, et al. Identifying specificity groups in the T cell receptor repertoire. Nature 2017; 547: 94–8.

2 Han A, Glanville J, Hansmann L, Davis MM. Linking T-cell receptor sequence to functional phenotype at the single-cell level. Nat Biotechnol 2014; 32: 684–92.

3 Halliday A, Whitworth H, Kottoor SH, et al. Stratification of Latent Mycobacterium tuberculosis Infection by Cellular Immune Profiling. J Infect Dis 2017; 215: 1480–1487.

4 Finak G, McDavid A, Chattopadhyay P et al. Mixture models for single-cell assays with applications to vaccine studies. Biostatistics. 2013;15:87–101.

